# Individual healthcare-seeking pathways for tuberculosis in Nigeria’s private sector during the COVID-19 pandemic

**DOI:** 10.1101/2023.06.13.23291334

**Authors:** Charity Oga-Omenka, Lauren Rosapep, Lavanya Huria, Nathaly Aguilera Vasquez, Bolanle Olusola-Faleye, Mohammad Abdullah Heel Kafi, Angelina Sassi, Chimdi Nwosu, Benjamin Johns, Abdu Adamu, Obioma Chijioke-Akaniro, Chukwuma Anyaike, Madhukar Pai

## Abstract

**Background:** Pre-COVID-19, individuals with TB in Nigeria were often underdiagnosed and untreated. Care for TB was mostly in the public sector while only 15% of new cases in 2019 were from the private sector. Reports highlighted challenges in accessing care in the private sector, which accounted for 67% of all initial care-seeking. Our study examined patients’ health seeking pathways for TB in Nigeria’s private sector, and explored any changes to care pathways during COVID, based on patients’ perspectives.

**Design/Methods:** We conducted 180 cross-sectional surveys and 20 in-depth interviews with individuals having chest symptoms attending 18 high volume private clinics and hospitals in Kano and Lagos States. Questions focused on sociodemographic characteristics, health-seeking behavior and pathways to care during the COVID-19 periods. All surveys and interviews were conducted in May 2021.

**Results:** Most participants were male (n=111, 62%), with average age of 37. Half (n=96, 53.4%) sought healthcare within a week of symptoms, while few (n=20, 11.1%) waited over 2 months. TB positive individuals had more health-seeking delays, and TB negative had more provider delays. On average, participants visited 2 providers in Kano and 1.69 in Lagos, with 61 (75%) in Kano and 48 (59%) in Lagos visiting other providers before the recruitment facility. Private providers were the initial encounters for most participants (n=60 or 66.7% in Kano, n-83 or 92.3% in Lagos). Most respondents (164 or 91%) experienced short-lived pandemic-related restrictions, particularly during the lockdowns, affecting access to transportation, and closed facilities.

**Conclusions:** This study showed a few challenges in accessing TB healthcare in Nigeria, necessitating continued investment in healthcare infrastructure and resources, particularly in the private sector. Understanding the different care pathways and delays in care provides opportunities for targeted interventions to improve deployment of services closer to where patients first seek care.

## Background

Before the COVID-19 pandemic, tuberculosis (TB) was the leading cause of infectious disease deaths globally [1]. Of the 10.6 million estimated cases in 2021 worldwide, only 6.4 million (60%) were diagnosed and notified [1]. Since the COVID-19 pandemic, global TB notifications and treatment have dropped, and deaths increased, erasing progress made in the last two decades [1]. This decline suggests increases in community transmission and infections.

Globally, several high burden TB countries reported that the COVID-19 pandemic and control measures significantly impacted case notifications, treatment, and other services for TB [1–3]. The End TB Strategies calls for early diagnosis and treatment of TB [4], yet delays are persistent in many high burden countries, including Nigeria, particularly in the private sector [5–8]. Extended delays are associated with increased risks of transmission, morbidity, and mortality [9–12].

Patient pathways analyses are useful approaches to understanding patients’ care seeking patterns and how they access available services [13, 14]. Individual patient pathway analyses can provide insights on the number of attempts individuals make to find TB care, and the missed opportunities for appropriate TB care. This information is necessary to surmount the problem of poor access to high quality TB care and is key to finding the missing millions of TB cases that go undiagnosed globally every year [1, 13, 15]. Pathway analyses have been used to highlight misalignment between demand and available resources at the macro level [14], or show the different care-seeking decisions at the micro level, as well as the types and numbers of providers that patients seek care with before getting diagnosed and treated [16, 17]. Several aspects of care pathways have been highlighted in literature, including time delays between symptom recognition, health seeking, diagnosis and treatment, as well as the number of healthcare encounters, type and sector of providers encountered, and point of diagnosis [14, 17, 18].

Nigeria, a high burden TB country, had an estimated 467,000 incident cases in 2021[1]. Case notification for TB in Nigeria has come mainly from the public sector. Only 15% of new TB cases in 2019 were reported from the private sector, even though it accounts for approximately 67% of initial TB care-seeking [19]. Nigeria recorded 88,388 confirmed cases of COVID-19 in 2020, with over 266,000 cases as of December 2022 [20]. During the initial lockdowns in March-April 2020, there were several reports indicating significant disruptions to TB services [21–24]. Nigeria and other high burden TB countries reported delays in care-seeking for respiratory symptoms due to fear of COVID-19 diagnosis [23, 25–28]. However, TB case finding began to rebound in June 2020 and by the second wave in December 2020 to March 2021, TB notifications had fully recovered to pre-pandemic levels [24]. This made Nigeria an outlier for pandemic recovery among other high-burden countries [1]. Private sector notifications significantly increased, contributing to the overall rise in TB notifications since 2020 [1, 29]. It remains unclear if there are long-lasting impacts of COVID-19 on healthcare seeking for chest symptoms in Nigeria’s private healthcare sector.

The USAID-funded *Strengthening Health Outcomes through the Private Sector* (SHOPS) Plus Nigeria project aimed to strengthen the private sector’s capacity to detect and treat TB in alignment with international standards and the national TB control plan. By training and supporting over 2,900 private providers, including clinical providers, laboratories, community pharmacists and proprietary patent medicine vendors (PPMVs) in Lagos and Kano States, the project contributed to substantial increases in private sector TB notifications between 2017 and 2020 [30]. Our study utilized cross sectional surveys and qualitative interviews with individuals with diagnosed and presumptive TB within the SHOPS Plus network in Kano and Lagos, to assess how healthcare seeking and pathways may have changed in the private sector. This study aimed to examine in Nigeria whether there were any long-lasting impacts of COVID on care-seeking given that the apparent disruptions in care were relatively short-lived.

### Conceptual framework

Our conceptual framework (*Fig 1*) is derived from existing literature [17, 31–33] and highlights two main types of delays along pathways to TB care: health-seeking and health system delays. Health-seeking delays refer to the time it takes from symptom onset to patients recognizing their symptoms and seeking care from a healthcare provider. Health-seeking is influenced by lack of TB knowledge, stigma, cost of seeking care, amongst other factors. Health system delays occur once patients have sought care and can be further divided into provider delays and treatment delays. Provider delays occur healthcare providers delay diagnosis due to low index of suspicion of TB, limited diagnostic tools, misdiagnosis, or delays in making referrals, conducting tests or transmitting results. Treatment delays occur when patients are diagnosed with TB, but there is a delay in initiating treatment, due to a lack of medication or equipment, insufficient staff, or poor communication between providers and patients. Diagnostic delays refer to the time it takes from the onset of symptoms (individual-level) to the confirmation of TB diagnosis (health-system level). Lack of knowledge of TB (symptoms, diagnosis and treatment) is a common factor contributing to diagnostic delay [5].

**Figure 1:**
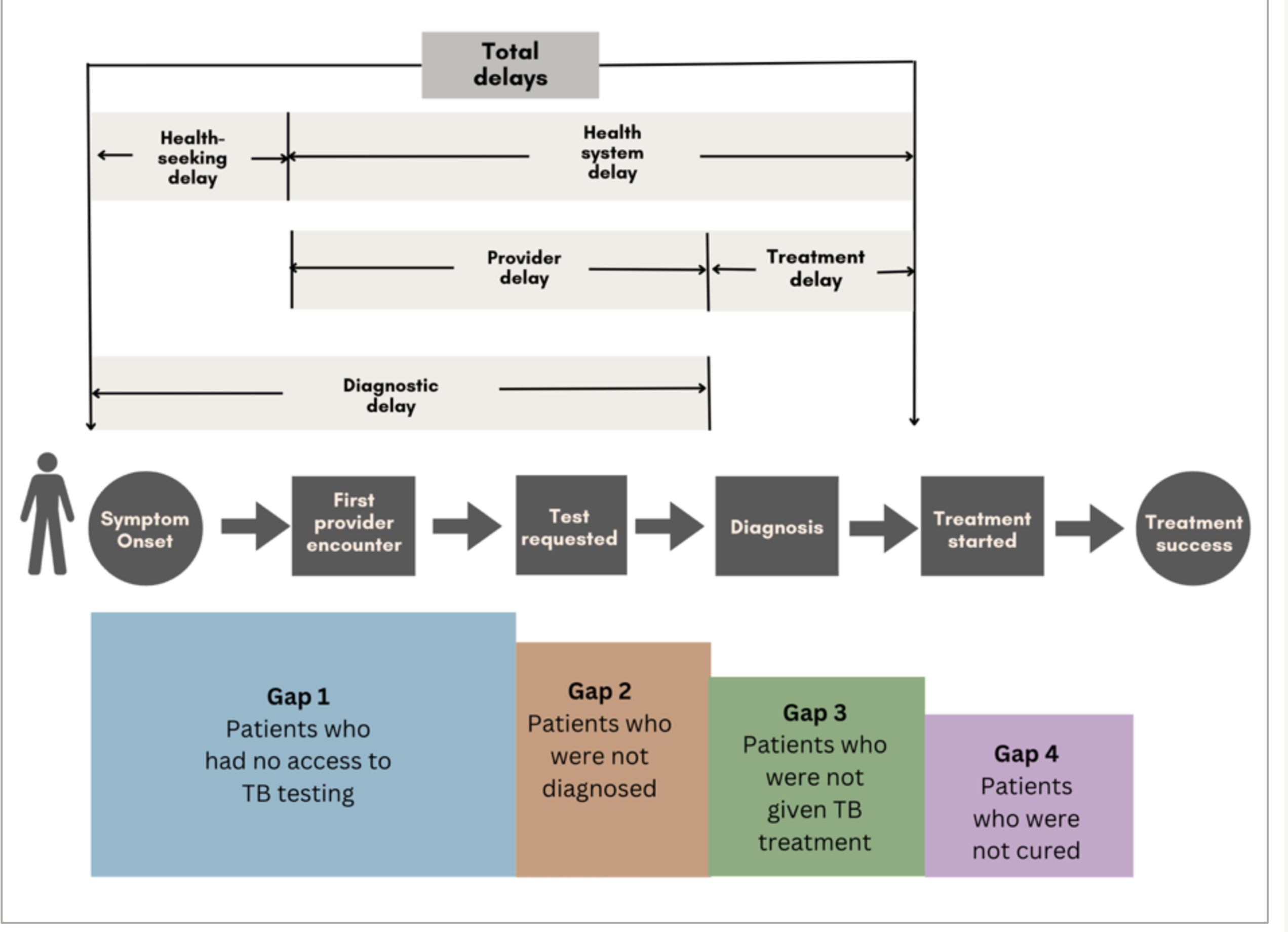
Conceptual framework on types of delays along the TB care cascade [17, 31-33]

There is some consensus that delays should not exceed 2-4 weeks from onset of symptoms to seek care, 3-7 days for testing (provider delay) using the current WHO-recommended TB diagnostics, and 1-2 days for treatment initiation to optimise patient outcomes [34–36]. The WHO considers as unethical any delay in treatment initiation in the presence of a positive diagnosis or a strong presumption of TB [35].

Previous studies conducted in Lagos [5] and rural Nigeria [7] found that health-seeking delays were more common than provider delays, contributing more to overall total delay. The factors associated with these delays can be categorized as individual-level (knowledge, attitudes, and behaviours), or structural-level (relating to healthcare facilities, providers, doctors, etc.).

These delays can lead to 3 major gaps or missed opportunities in accessing TB care: Gap 1 includes those who never access TB testing, Gap 2 are those who access testing but are not diagnosed, and Gap 3 includes those diagnosed but not treated. These gaps result in prolonged illness, disease transmission, increased risk of drug-resistant TB, and death [1, 32, 33]. Studies done in Nigeria before the pandemic identified Gap 1 as the biggest challenge in TB control, followed by Gap 2 [6, 32, 37, 38].

## Methods

### Study design, population and sampling

We conducted pathway analyses using mixed methods, to survey 180 individuals and conduct 20 in-depth interviews during the second COVID-19 wave in May 2021 (*Appendix 1*). This study was part of a larger research project in 3 high burden countries. We previously published the timelines of the COVID epi-curve, TB notifications and the timing of our data collection in Nigeria [24]. Our study took place among individuals seeking TB care in 18 private facilities in Kano and Lagos, the states with the highest TB burden and levels of private sector activity [30], and with estimated population of 14.7 million and 13 million respectively in 2021 [39]. We sampled active providers within the SHOPS Plus program network, 1,084 in Kano and 1,317 in Lagos, out of which were 217 and 472 clinical facilities respectively.

To ensure even geographic distribution of recruitment facilities within the states, we used a non-probabilistic quota sampling to select 18 high volume TB clinical facilities within the SHOPS Plus network (*Appendix 2*). We prioritized higher volume facilities to minimize overall facility and network burden and increase recruitment efficiency. We included the three highest volume facilities (by total TB notifications) in each senatorial zone, ensuring only one facility per local government areas (LGA), resulting in a total of nine facilities per state.

We visited each consenting recruitment facility and developed a list of eligible patients from the facility’s National TB program mandated presumptive and treatment registers, from which participants were randomly selected. Two types of patients were recruited – patients confirmed to be negative for TB (negative sputum test result in the register); and patients recently diagnosed (positive sputum test result for TB) receiving or scheduled for TB treatment within the same facility. All recruited patients were over 18 years old, HIV negative, and had confirmed results between January and May 2021 (to minimize survey recall bias). We excluded previously confirmed COVID-19 cases, individuals unable to provide a sputum sample, patients initiating treatment at another facility, and those with missing sputum results or a history of anti-TB treatment in the past 6 months. Within each recruitment facility, we surveyed ten participants and conducted additional in-depth interviews 1-2 participants.

Eligible individuals were contacted by facility staff to explain the study’s purpose and assess their initial willingness to participate. A date, time and place (the clinical facility, patient’s home, or place of work) were agreed upon for the survey and interviews.

### Data collection

Our study instruments focused on various aspects, including the onset of symptoms and care-seeking pathway, interactions with healthcare providers from initial encounters to TB diagnosis and treatment initiation, healthcare decision-making and use during COVID-19, perspectives on the pandemic, and the impact of COVID-19 on patient care.

To ensure accuracy, the study tools were translated into local languages (Yoruba and Hausa), back translated to English, pretested and piloted. Survey instruments were programmed into SurveyCTO and field officers were trained for each survey, and only those who performed satisfactorily were deployed. Precautionary measures against COVID-19 were implemented prior to survey implementation, and verbal informed consent was obtained before interviews. Transport costs were reimbursed for participants who preferred interviews in the facility. Completed tools were uploaded to the SurveyCTO server. We maximally varied interview participants by location, TB status, and gender.

Interviews, which lasted 24 to 55 minutes on average, were conducted in English, with respondents encouraged to request translation into or respond in Nigerian Pidgin, Yoruba or Hausa if needed. All interviews were audio-recorded and transcribed, with translations from Yoruba and Hausa languages by fluent translators.

### Data analysis and pathway construction

Our analysis involved descriptive statistics, construction of care pathways and logistic regressions. We weighted the data to address non-response and ensure the similarity between responders and non-responders in terms of background characteristics [40, 41]. We used logistic regression with covariates to calculate non-response weight, and predictive probabilities of survey response were calculated derived. Unweighted mean of probabilities matched the 70% unweighted response rate in dataset. [3]. The non-response weight was determined by inverting the predictive probabilities [41], resulting in a sum equal to the sample size.

We used descriptive statistics to analyze demographic characteristics of non-responders, considering both weighted and unweighted data. Univariate Pearson’s chi-square (Χ2) tests assessed associations between individual characteristics, TB status, private sector TB care delivery, impact of COVID, using weighted and unweighted cases.

We constructed individual patient-level pathways using stacked bar charts, which depict the chronological order in which individuals encountered various types of healthcare providers leading up to diagnosis and treatment. To create these sequencing charts, we accounted for points of diagnosis (including laboratories, community outreaches and traditional healers) even where the participants themselves did not acknowledge these as unique provider encounters.

Logistic regressions (weighted) were utilized to examine the relationship between patients’ characteristics and symptoms with their utilisation of private providers as the initial point of seeking healthcare, the number of provider encounters, as well as the use of multiple providers. We reported the odds ratio with 95% confidence intervals, considering a p-value of < 0.05 as statistically significant.

Statistical analysis was performed using the open-source programming language R (version 4.0.3) and R Studio (version 1.4.1106). Transcripts were coded inductively and deductively by two coauthors, CO and LH, using the Quirkos software (version 1.61), with an initial codebook developed from 2 transcripts. Codes and themes were double-checked by CO, and the thematic analysis reviewed by the research team for alignment with study objectives.

### Ethical considerations

Our study received ethical approval from the McGill University Health Centre (MUHC) Research Ethics Board (REB), with approval number Covid BMGF / 2021-7197. We also obtained approval from the health research ethics committee (HREC) in both States, including from the Lagos State University Teaching Hospital (LASUTH) and Kano State Ministry of Health (MoH). We also obtained letters of introduction from Abt Associates to the MoH, National Association of Patent and Proprietary Medicines (NAPPMED), Association of Community Pharmacists in Nigeria (ACPN), and heads of the target facilities. All survey and interview participants gave written or verbal consent.

## Results

### Characteristics of participants and non-response

Out of 337 patients contacted (*Appendix 3*), 180 participants completed the survey and 20 participated in the in-depth interviews. Higher non-response rates were observed among participants from Kano State (62.5%) and those who tested negative for TB (54%). Gender and age distributions were similar for both surveyed and non-surveyed participants, except for the 55+ age group, where non-surveyed rate was higher (18.8% vs 12.2%).

The participants (*Table 1*) were predominantly male (62%), married (53%) especially in Kano State (58 vs 49% in Lagos), with an average age of 37 years. Participants had in Kano had more dependent relatives on average (6) than in Lagos (3). Larger proportions were self-employed (43%), had a secondary school education (34%). More participants lived within a 2km radius from a public facility (48%), or a private facility (54%).

**Table 1:** Characteristics of surveyed participants by States [N = 180]

|  | KANO | LAGOS |
| --- | --- | --- |
| Gender |  |  |
| Male | 60 (66.7%) | 51 (56.7%) |
| Female | 30 (33.3%) | 39 (43.3%) |
| Age |  |  |
| Mean (SD) | 35.9 (13.8) | 38.5 (12.9) |
| TB status |  |  |
| TB negative | 45 (50.0%) | 46 (51.1%) |
| TB positive | 45 (50.0%) | 44 (48.9%) |
| Marital status |  |  |
| Married | 52 (57.8%) | 44 (48.9%) |
| Single | 33 (36.7%) | 36 (40.0%) |
| Widowed | 2 (2.2%) | 6 (6.7%) |
| Separated | 0 (0%) | 4 (4.4%) |
| Divorced | 3 (3.3%) | 0 (0%) |
| Number of dependent relatives |  |  |
| Mean (SD) | 5.77 (4.49) | 2.77 (2.02) |
| Median [Min, Max] | 5.00 [0, 28.0] | 3.00 [0, 11.0] |
| Highest education |  |  |
| Senior secondary | 29 (32.2%) | 32 (35.6%) |
| Primary 6 or lower | 9 (10.0%) | 20 (22.2%) |
| Bachelor's or higher | 7 (7.8%) | 15 (16.7%) |
| OND/HND* | 12 (13.3%) | 5 (5.6%) |
| Junior secondary | 3 (3.3%) | 8 (8.9%) |
| Trade or vocational | 2 (2.2%) | 4 (4.4%) |
| Missing | 28 (31.1%) | 6 (6.7%) |
| Current employment |  |  |
| Self-employed | 45 (50.0%) | 33 (36.7%) |
| Employee | 19 (21.1%) | 41 (45.6%) |
| Not working | 26 (28.9%) | 16 (17.8%) |
| Nearest public facility |  |  |
| Less than 2 km | 55 (61.1%) | 31 (34.4%) |
| Between 2-10 km | 32 (35.6%) | 27 (30.0%) |
| Don't know/not sure | 1 (1.1%) | 19 (21.1%) |
| Over 10km | 2 (2.2%) | 13 (14.4%) |

*\*OND/HND Ordinary National Diploma/Higher National Diploma (representing 2-3 years post-secondary education)*

### Health-seeking and health system delays

Half of the 180 participants (n=96, 53.4%) sought care with a provider within a week of noticing their symptoms, with 17.8% seeking care within the first 2 days *(Table 2)*. Only very few patients (n=20, 11.1%) waited for 2 months and above before seeking care.

**Table 2:** Delays, care pathway, missed opportunities and the impact of COVID-19

|  | Unweighted |  | Weighted |  | p-value |
| --- | --- | --- | --- | --- | --- |
|  | KANO | LAGOS | KANO | LAGOS |  |
| **Delay between symptom onset and first provider encounter** |  |  |  |  |  |
| 1 or 2 days | 12 (13.3%) | 20 (22.2%) | 12.8 (13.5) | 18.9 (22.6) | 0.102 |
| 3 to 7 days | 37 (41.1%) | 27 (30.0%) | 39.5 (41.6) | 25.4 (30.3) |  |
| 1 to 4 weeks | 21 (23.3%) | 17 (18.9%) | 22.0 (23.2) | 15.8 (18.9) |  |
| 1 to 2 months | 10 (11.1%) | 16 (17.8%) | 10.2 (10.8) | 14.7 (17.57) |  |
| 2 to 3 months | 5 (5.6%) | 1 (1.1%) | 5.2 (5.5) | 0.9 (1.12) |  |
| Over 3 months | 5 (5.6%) | 9 (10.0%) | 5.2 (5.5) | 8.0 (9.58) |  |
| **Sought care at more than one facility** |  |  |  |  |  |
| Yes | 61 (67.8%) | 48 (53.3%) | 63.8 (67.3%) | 44.0 (52.6%) | 0.048 |
| No | 29 (32.2%) | 42 (46.7%) | 31.1 (32.7%) | 39.7 (47.4%) |  |
| **Number of encounters +** |  |  |  |  |  |
| Mean (SD) | 2 (0.936) | 1.69 (0.76) | 2.0 (0.9) | 1.7 (0.75) | 0.015 |
| Median [Min, Max] | 2 [1, 5] | 2 [1, 4] | 2 [1, 5] | 2 [1, 4] |  |
| **Number of providers encountered +** |  |  |  |  |  |
| Only at recruitment facility | 29 (25%) | 42 (41%) | 31.1 (32.7%) | 39.7 (47.4%) | 0.183 |
| Two provider encounters | 58 (50%) | 46 (45%) | 63.8 (67.3%) | 44.0 (52.6%) |  |
| Three provider encounters | 21 (18%) | 12 (12%) | 21.7 (22.9%) | 10.8 (12.9%) |  |
| Four provider encounters | 6 (5%) | 2 (2%) | 6.1 (6.4%) | 1.8 (2.1%) |  |
| Five provider encounters | 2 (2%) | 0 (0%) | 2.0 (2.1%) | 0 |  |
| **First provider contacted after symptoms** |  |  |  |  |  |
| Private provider | 60 (66.7%) | 83 (92.3%) | 63.3 (66.7%) | 77.5 (92.5%) | 0.001 |
| Public provider | 27 (30.0%) | 5 (5.5%) | 28.2 (29.4%) | 4.5 (5.4%) |  |
| Informal provider | 22 (33.3%) | 2 (42.2%) | 3.3 (3.5%) | 0.9 (1.1%) |  |
| **Testing with first provider contacted** |  |  |  |  |  |
| Provider did not discuss COVID | 55 (61.1%) | 40 (44.4%) | 57.3 (60.4%) | 36.8 (44.0%) | 0.208 |
| Provider did not discuss TB* | 39 (43.3%) | 32 (35.6%) | 40.4 (42.6%) | 29.4 (35.1%) | 0.154 |
| Provider said lab test was needed* | 36 (40.0%) | 22 (24.4%) | 37.7 (39.7%) | 20.1 (23.9%) | 0.055 |
| Provider recommended sputum test* | 25 (27.8%) | 14 (15.6%) | 26.7 (28.1%) | 12.8 (15.3%) | 0.067 |
| **Time from TB diagnosis to treatment start** |  |  |  |  |  |
| Same day or within 2 days | 34 (37.8%) | 32 (35.6%) | 34.1 (60.4%) | 28.8 (32.0%) | 0.075 |
| 3-7 days | 3 (3.3%) | 10 (11.1%) | 3.2 (3.5%) | 9.1 (10.1%) |  |
| 8- 30 days | 4 (4.4%) | 2 (2.2%) | 4.1 (4.6%) | 1.9 (2.1%) |  |
| 1- 2 months | 2 (2.2%) | 0 (0%) | 1.9 (2.1%) | 0 (0%) |  |
| Over 2 months | 2 (2.2%) | 0 (0%) | 2.2 (2.4%) | 0 (0%) |  |
| Negative for TB | 45 (50.0%) | 46 (51.1%) | 49.4 (54.9%) | 43.9 (48.8%) |  |
| **Impact of COVID-19 the health facility most frequented (multichoice)** |  |  |  |  |  |
| New infection control measures | 46 (51.1%) | 54 (60.0%) | 48.7 (51.3%) | 50.6 (60.4%) | 0.222 |
| Reduced hours | 16 (17.8%) | 3 (3.3%) | 16.9 (17.8%) | 2.69 (3.2%) | 0.001 |
| Screening for COVID-19 | 8 (8.9%) | 4 (4.4%) | 8.5 (8.9%) | 3.63 (4.3%) | 0.213 |
| Closed | 3 (3.3%) | 2 (2.2%) | 3.2 (3.4%) | 1.81 (2.2%) | 0.617 |
| **Since the pandemic in March 2020, has there been any time when you needed health care but did not receive it?** |  |  |  |  |  |
| No/ Cannot remember | 84 (93.3%) | 88 (97.8%) | 88.4 (93.1%) | 81.87 (97.8%) | 0.130 |
| Yes | 6 (6.7%) | 2 (2.2%) | 6.5 (6.9%) | 1.81 (2.2%) |  |
| **How COVID-related restrictions affected access to healthcare since March 2020 (multichoice)** |  |  |  |  |  |
| There has been no change for me. | 57 (63.3%) | 57 (63.3%) | 59.8 (63.0%) | 52.79 (63.1%) | 0.996 |
| Unable to leave the house to seek care because of movement/transport restrictions | 20 (22.2%) | 21 (23.3%) | 21.1 (22.2%) | 19.97 (23.9%) | 0.792 |
| Unable to reach a doctor because facilities were closed | 15 (16.7%) | 12 (13.3%) | 15.7 (16.5%) | 11.28 (13.5%) | 0.574 |
| Waiting times at facilities were longer than normal/expected | 15 (16.7%) | 12 (13.3%) | 16.3 (17.1%) | 11.35 (13.6%) | 0.513 |
| Unable to receive my medications because drug stores were closed | 9 (10.0%) | 6 (6.7%) | 9.6 (10.1%) | 5.72 (6.8%) | 0.439 |
| Sought care from a different facility than typical/preferred facility | 8 (8.9%) | 3 (3.3%) | 8.5 (8.9%) | 2.85 (3.4%) | 0.130 |
| **How has COVID and/or current/past COVID-related restrictions affected <u>your willingness</u> to seek healthcare?** |  |  |  |  |  |
| I am more willing to seek care | 52 (57.8%) | 66 (73.3%) | 55.0 (57.9%) | 61.02 (72.9%) | 0.001 |
| No effect | 38 (42.2%) | 18 (20%) | 39.9 (42.0%) | 17.13 (20.5%) |  |
| Don't know/Not sure | 2 (2.2%) | 1 (1.1%) |  |  |  |
| I am less willing to seek care | 0 (0%) | 6 (6.7%) | 0 | 5.54 (6.6%) |  |

\* These rates are calculated across as they form part of Yes or No questions in the survey tool. Participants who answered 'Yes' to the question are shown in the table, as a fraction of total respondents to that question.
+ The number of encounters used in the tables and regression were self-reported by the participants

Health-seeking and provider delays were experienced more by participants diagnosed with TB (38% and 43% *Fig 2a*), with 14% of them experiencing treatment delays. Participants from Lagos (29%, *Fig 2b*) experienced more health-seeking delays. Provider and treatment delays were more in Kano (40% and 10%) compared to in Lagos (28% and 4%).

**Figure 2:**
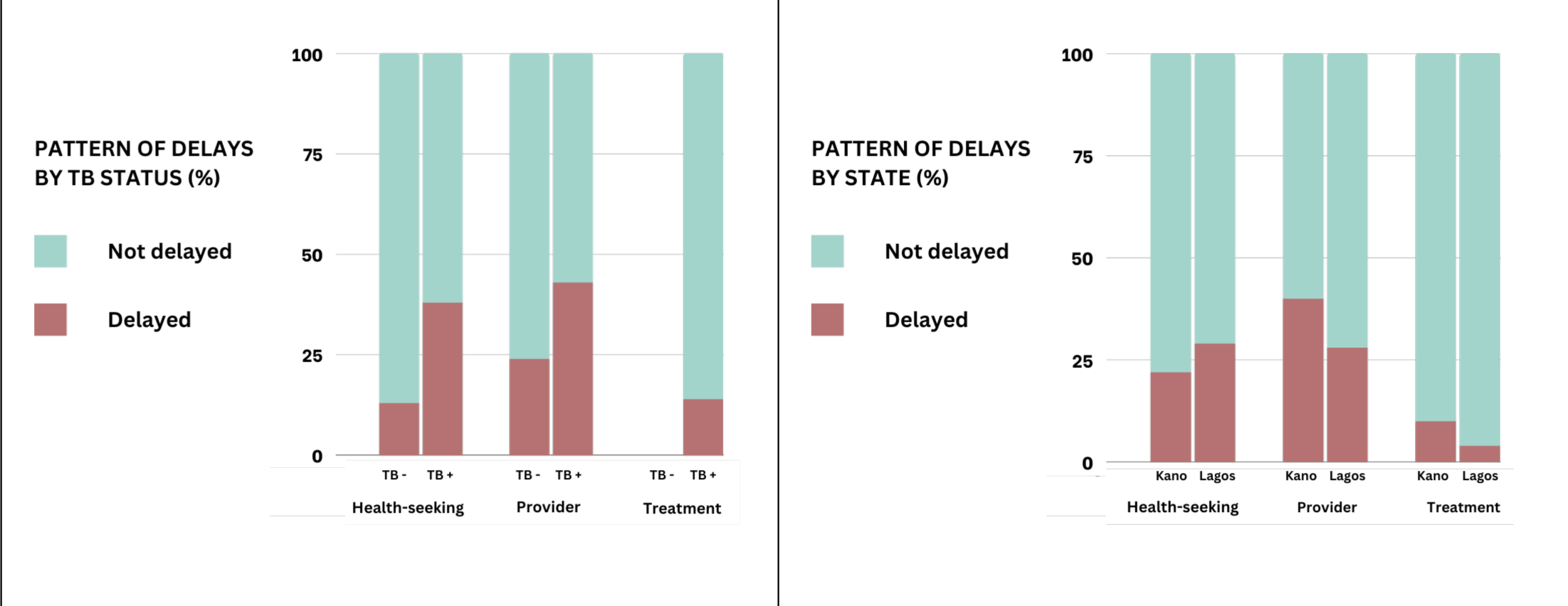
The proportion of participants experiencing different types of delays (a) by TB status and (b) by State

Out of 180 participants, (n=71, 39%) had their sole provider encounter at the study recruitment facilities (private clinics or hospitals), which included 3 individuals who had previously been diagnosed at community outreaches (*Table 2*). *Fig 2c* presents the number of participants and the varying time delays at each provider encounter. Among 109 participants who visited other providers before the recruitment facility, 55 had their initial encounter within a week, 25 waited 1-4 weeks, and 29 waited over a month. Out of the 33 who consulted two additional providers, 8 waited 1-7 days from the first encounter, 16 waited 1-4 weeks, and 9 waited for over a month. Among 8 who visited 3 providers, 4 waited 1-7 days following the second encounter, 1 waited 1-4 weeks, and 3 waited for more than a month. Of the 2 who visited 4 providers, 1 waited 1-7 days, and the other waited 1-2 months.

**Figure 2c:**
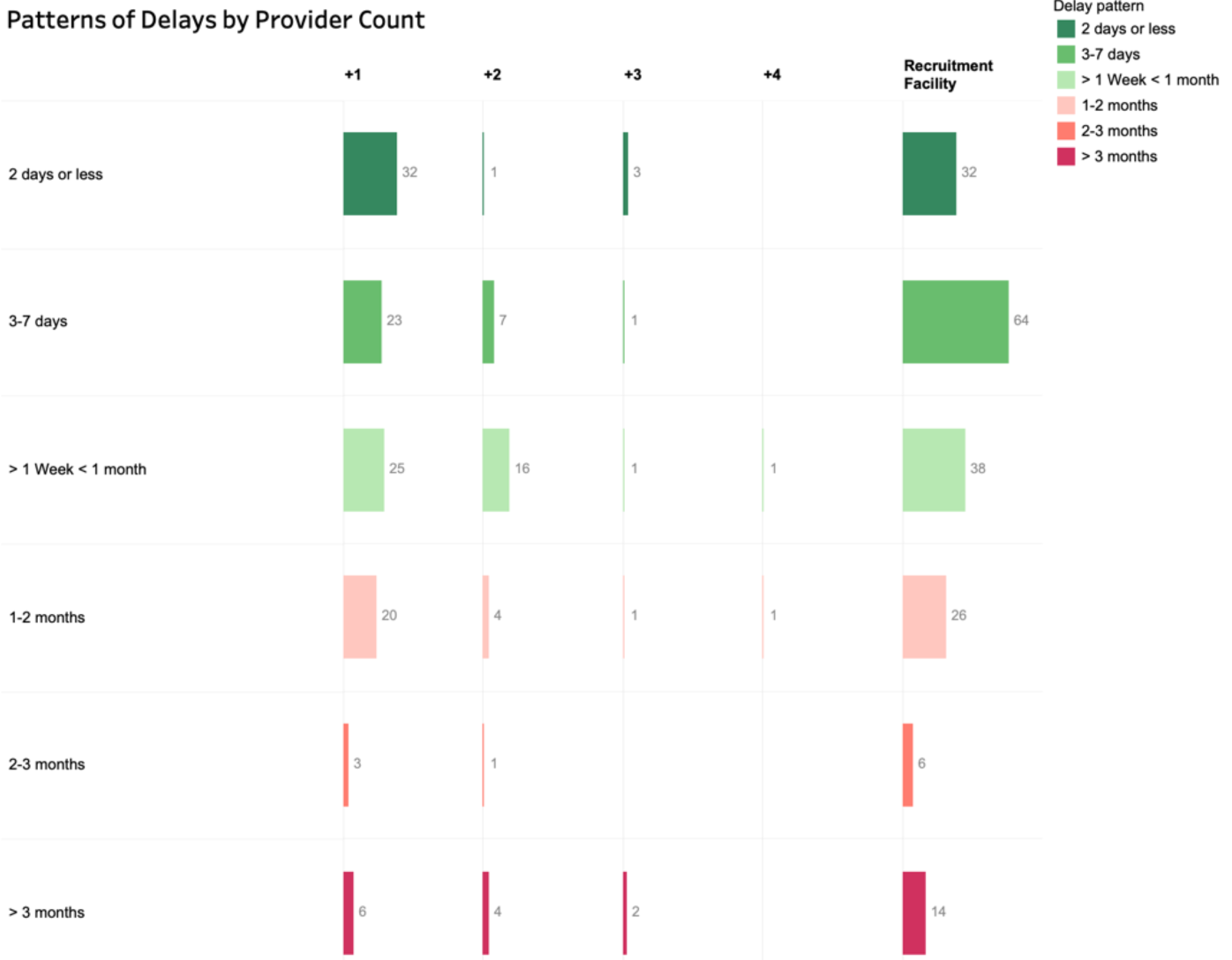
Patterns of delays with each additional provider encounter for 180 participants

At the recruitment facility, 96 of 180 participants waited 1-7days, 38 waited 1-4 weeks and 46 waited more than 1 month. Of those who waited more than a month, the majority were individuals who had visited other providers. Delays between encounters increase with each additional provider encounter, particularly with the 3^rd^ and 4^th^ encounters. Most TB patients were initiated on treatment within 2 days of diagnosis (n=34, 75% in Kano; n=32, 73% in Lagos).

From the qualitative interviews, many participants, regardless of TB status, reported health-seeking delays as they waited until their symptoms became prolonged. Symptom minimisation was a very common theme as most participants did not take coughing to be a serious health issue, at first (*Fig 3*). Participants assumed that the cough would subside, and participants self-medicated. Some participants mentioned that they were also afraid of getting infected with or being diagnosed with COVID-19. Provider delays were mostly due to misdiagnosis and delayed referral to TB testing sites. All TB patients interviewed in Kano and Lagos were initiated on treatment within 2 days of diagnosis.

**Figure 3:**
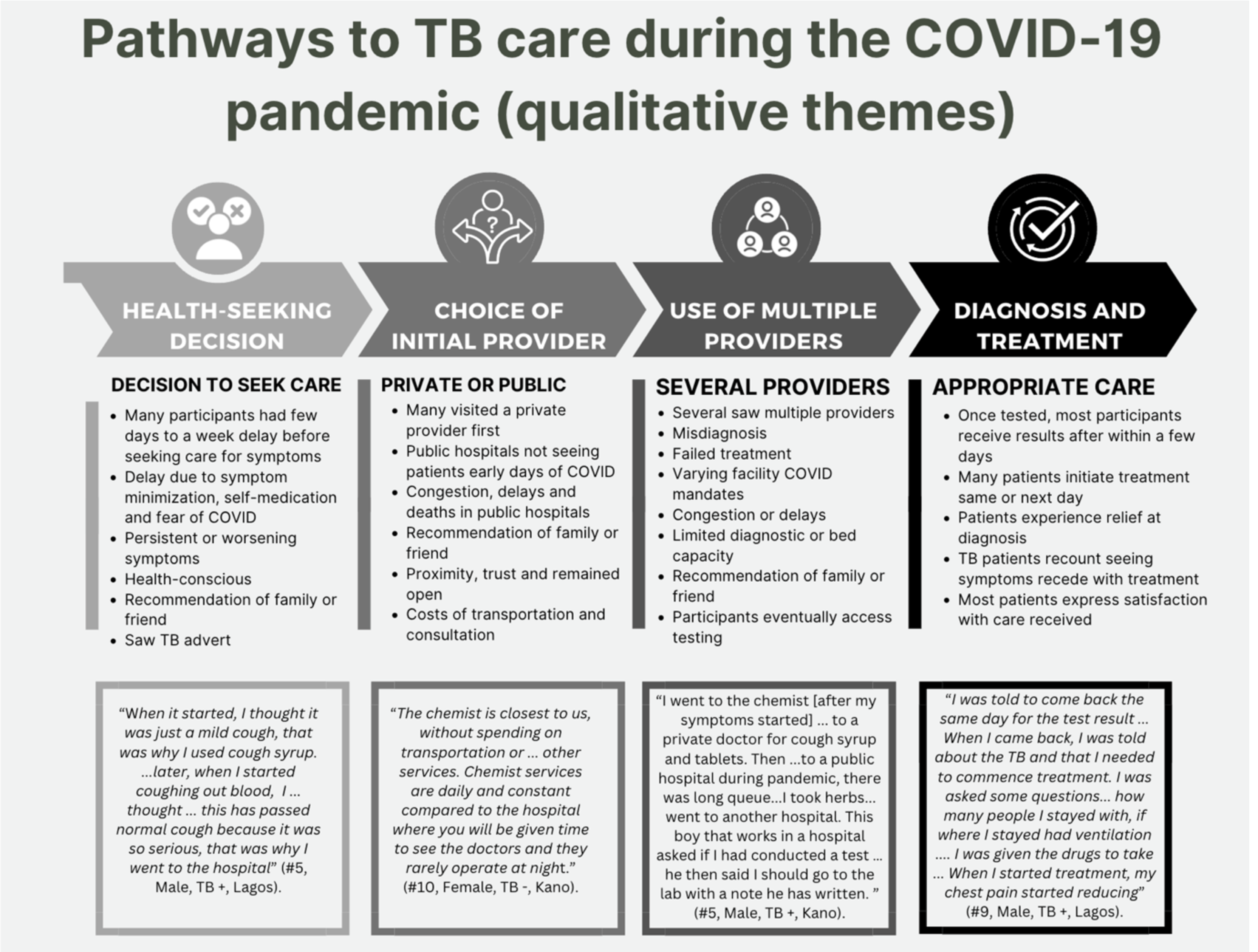
Qualitative themes on pathways to TB care

### Individual patient pathways

On average, participants had 2 provider encounters in Kano, and 1.69 in Lagos, with a median of 2 in both States before visiting the recruitment facility *(Appendix 4)*. Participants who visited the network of private hospitals and clinics had the fewest average encounters (mean of 1.10) compared to other providers (pharmacies or vendors = 2.35, public clinic = 3.00, traditional healers = 3.00, private clinic=3.33). Our survey revealed that the majority (n=109, 61%) patients sought care from multiple providers. Twenty-five percent of participants in Kano, and 41% in Lagos sought care only in the recruitment facility.

Our pathway sequencing charts (*Fig 4a and 4b*) identified 31 and 16 different patterns in Kano and Lagos, respectively, of progressive provider encounters. Participants in Kano, irrespective of their diagnosis, had more encounters and more variation in health-seeking than in Lagos. The largest share of participants in Kano used one additional provider (n=42, 46.6%) while for Lagos, the majority (n=42, 46.6%) used the recruitment facility (RF) exclusively.

**Fig 4a and 4b:**
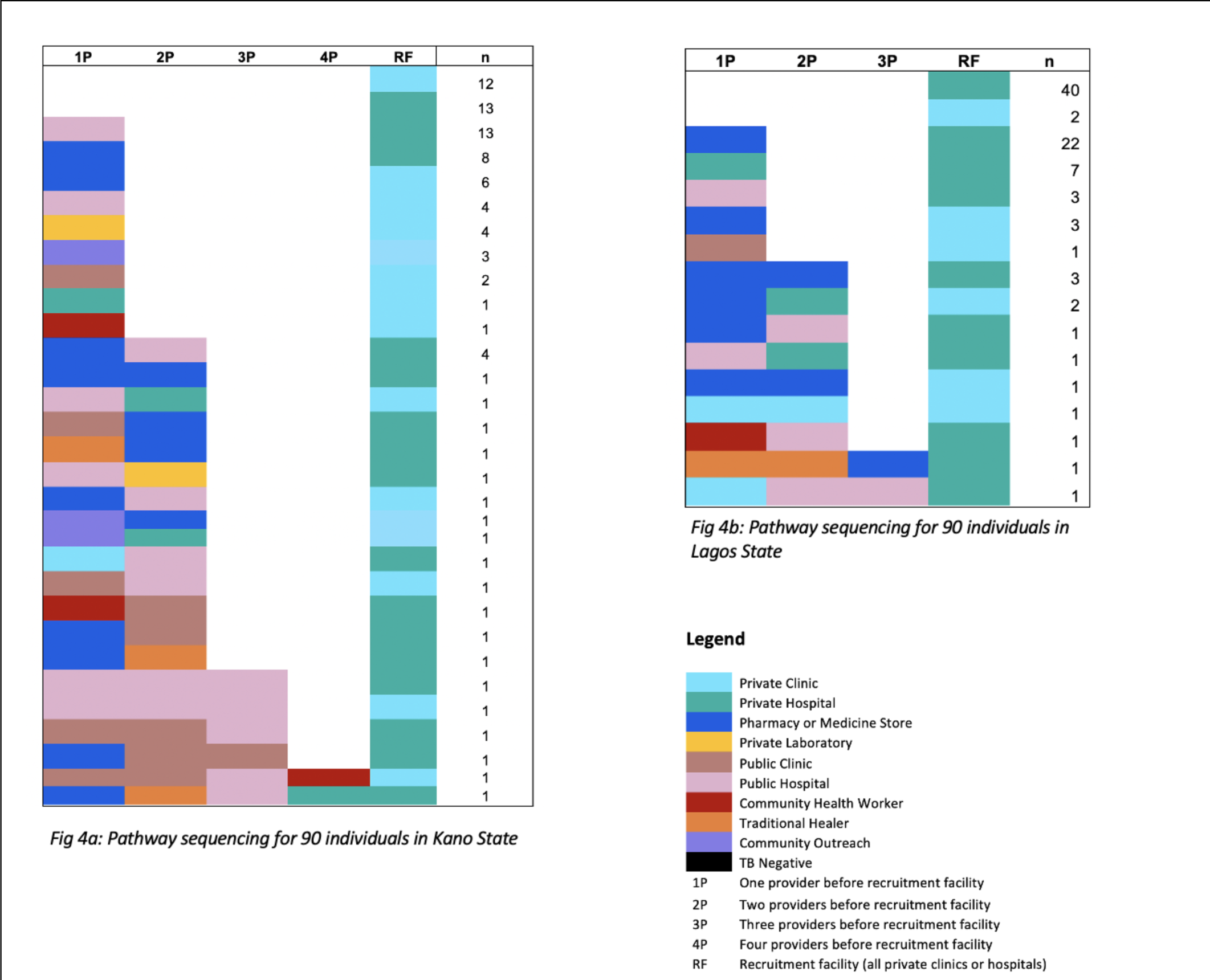
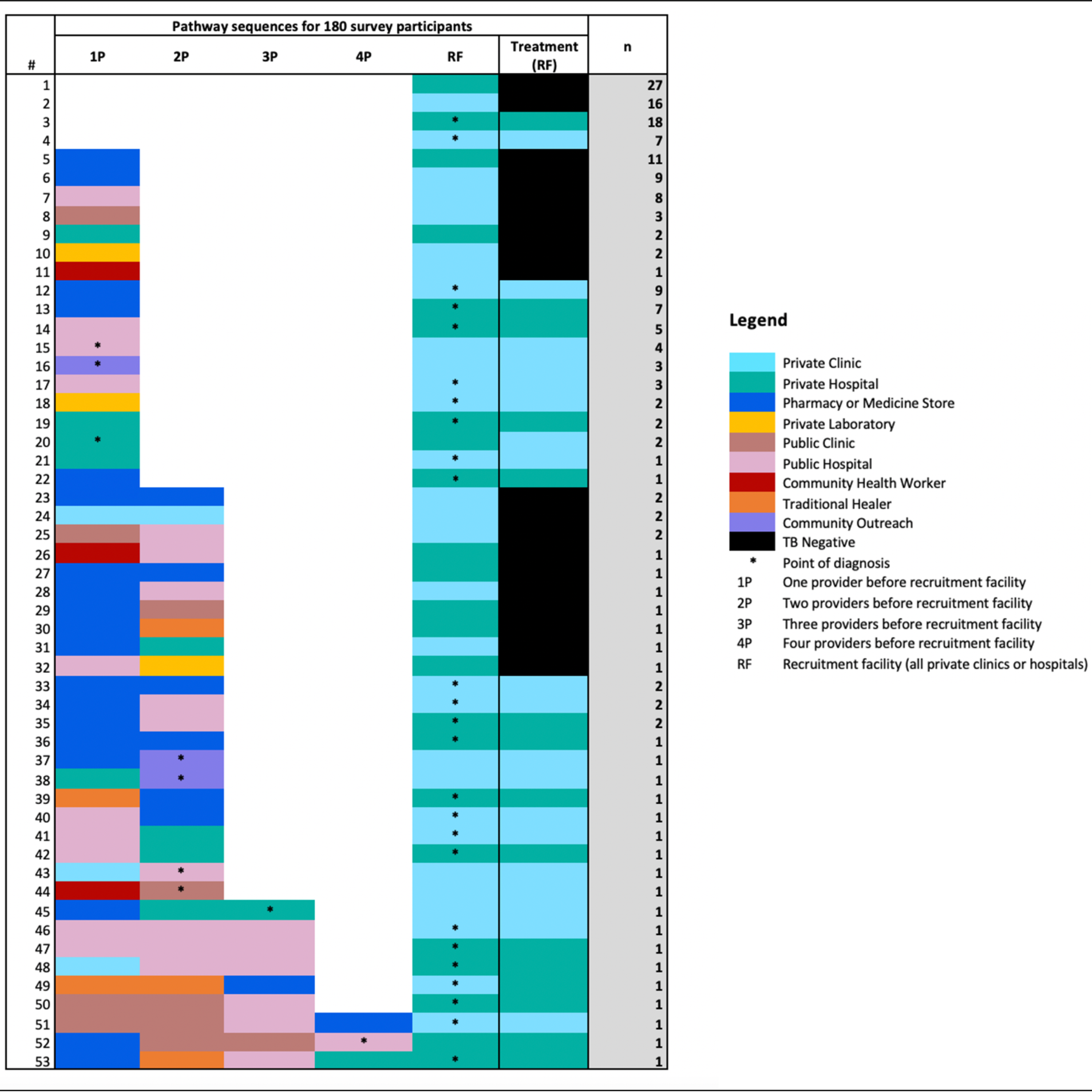
Pathway sequences for all survey participants and those in each state

*Fig 4c* showed 53 different sequencing patterns for all 180 survey participants, and included points of diagnosis, as well as the proportions who ended up with a negative diagnosis. The majority of participants (n=165, 91.6%) were diagnosed (with or without TB) at the recruitment facilities. All diagnosis points were clinical facilities (private or public). All TB positive participants (n=90) were initiated on treatment in the private hospitals or private clinics that were our recruitment facilities.

The qualitative data (*Fig 3, Appendix 5*) shows many participants used a private provider first, because these providers were more accessible and convenient, and public hospitals were congested. In terms of choice of provider, this was either because of proximity, prior relationship with provider, cost considerations or due to referral or contact tracing from someone the participant knew. Several of those with more than one encounter said that the medicine store was their first port of call. Many participants visited multiple providers as their symptoms worsened, particularly those with TB. For participants diagnosed, once they were told they had TB, several said they were placed on treatment on the same or next day. Although the pathways to diagnosis were cumbersome for many, they expressed satisfaction with the care they received afterwards. The majority of themes regarding treatment were positive experiences after diagnosis.

### Impact of COVID-19

At the time of our survey, 74 (41%) participants said it was easier to access healthcare, while 46 (26%) said it was still difficult. Most respondents (164 or 91%) experienced lockdowns or other pandemic-related restrictions, which affected transportation to seek care for 41 (23%), closed facilities for 27 (15%) and longer waiting times for 27 (15%) of them. However, at the time of the survey, most respondents said they were more willing (118 or 66%) to seek healthcare, compared to before the pandemic.

From the in-depth interviews, early on in the pandemic, participants were concerned about the rising deaths due to COVID-19, which discouraged them from going to a clinic or hospital. Participants didn’t want to get infected, but more importantly, they didn’t want to be forced to take a diagnostic test for COVID-19, test positive, then have to quarantine, which was the practice in many public hospitals at the time. A few participants said transport challenges during the lockdown period hampered their access to care. Participants also mentioned provider attitudes during the early phase of the pandemic as being fearful or unhelpful.

### Determinants and factors influencing private sector use and number of encounters

We looked at predictors for two outcome variables – 1) Choosing to use a private facility as their first healthcare contact 2) Using more than one provider from symptom onset. We also present the adjusted regression coefficient for the number of provider encounters.

When we looked at the predictors for participants choosing to use a private facility as their first healthcare contact (*Fig 5a*), we found that participants in Lagos State were more likely (OR = 8.97, 95% CI: 2.90 – 27.65). Those who were self-employed (OR = 0.22, 95% CI: 0.06 – 0.76), and those presenting with rashes and allergies (OR = 0.16, 95% CI: 0.03 – 0.79), were less likely to use private providers first. No other variables, including age, gender, TB status or symptoms were significant. The only statistically significant determinant of using more than one provider (*Fig 5b*) was having difficulty in breathing as a symptom (OR = 4.69, 95% CI: 1.42 – 15.51).

**Figure 5a:**
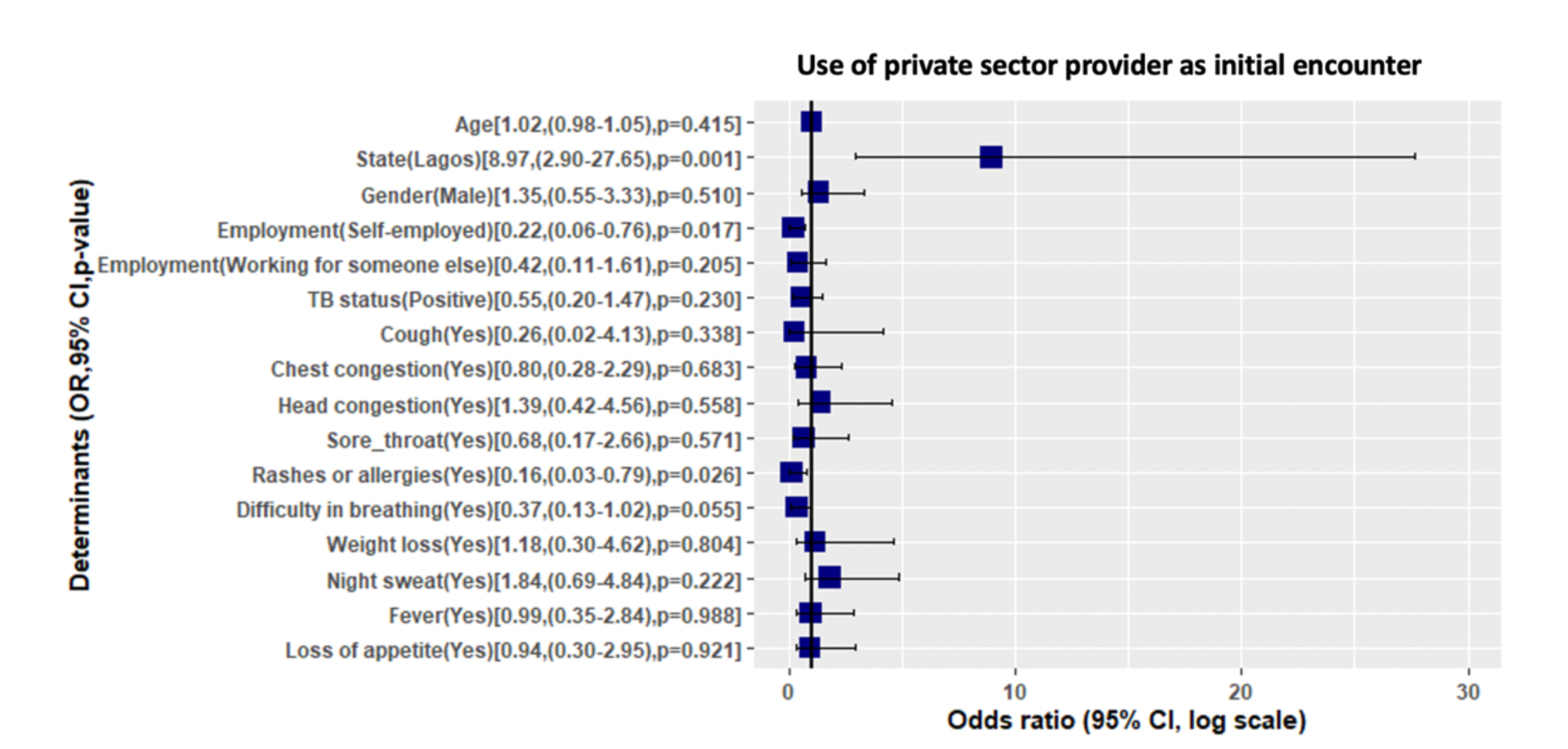
Determinants of using the private sector as first provider contact

**Figure 5b:**
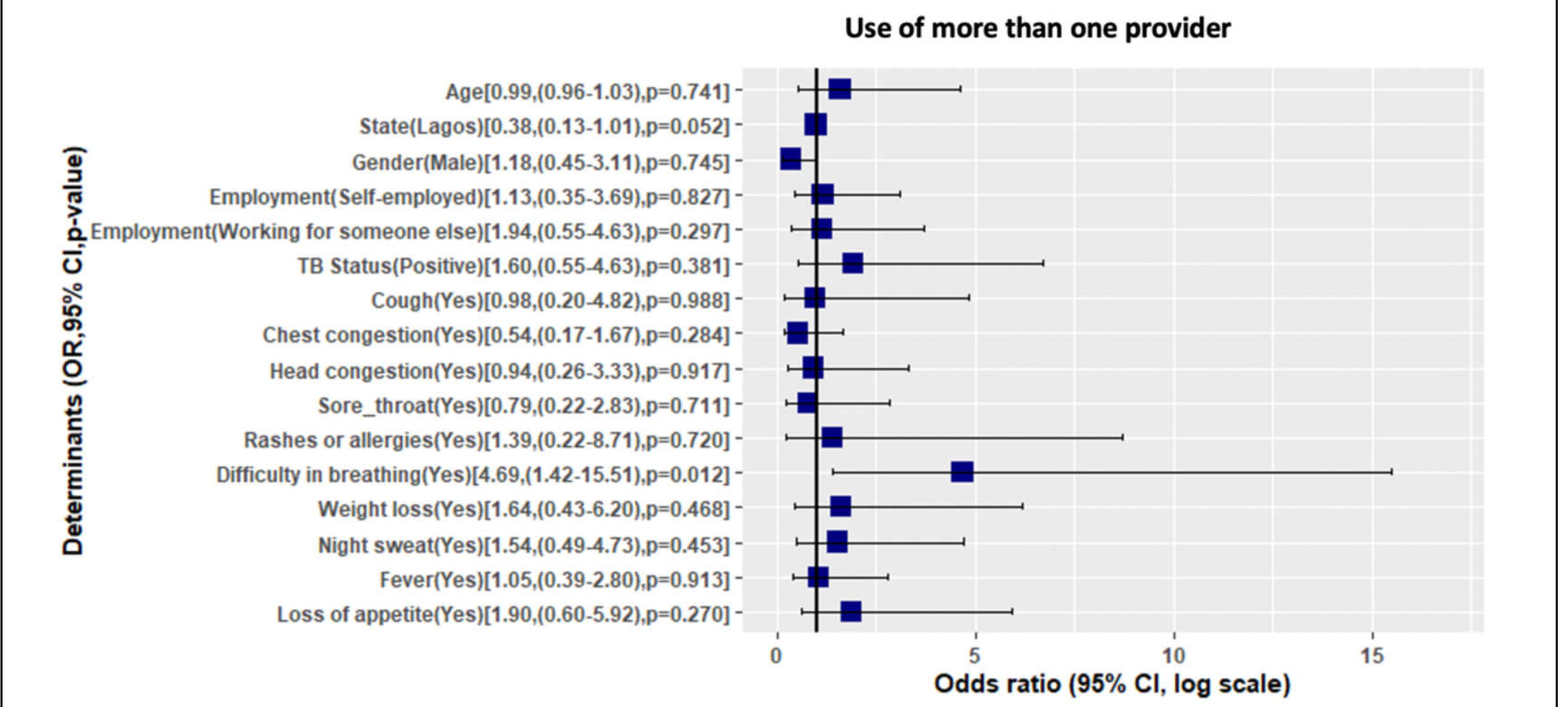
Determinants of using more than one provider after symptoms

## Discussion

In our study of patient pathways within the private sector in two Nigerian states, we found that individuals with chest symptoms experienced relatively short pathways. The study was conducted in a unique context, differing from the typical urban areas in Nigeria. The sampled providers belonged within the SHOPS Plus network, which provided extensive support to private sector facilities, ensuring efficient referral systems, availability of laboratory equipment and medications, and community engagement for case identification. Even within this context, patients faced some health-seeking and provider delays.

### Health-seeking and health system delays

Our investigation showed several challenges associated with health-seeking and provider delays. However, once individuals with TB were identified and given a definitive diagnosis, their experience with the healthcare provider was much more positive and timelier than their experience before diagnosis, agreeing with several studies in Nigeria and other high burden settings [32, 38, 42]. Of the 90 TB positive participants, majority (n=66, 73%) and all of the qualitative interview participants were treated within 2 days of diagnosis, contributing to their feelings of relief or gratitude, because they finally felt, ‘seen’ by the healthcare system.

### Individual pathways and private sector use

Among the 180 participants and excluding the 71 who sought care first at the recruitment facility, the majority (74 out of 109) initially sought care in private facilities. Some studies in Nigeria and in similar high burden TB countries have found that patients seek care first in drug stores or with traditional healers before clinics or hospitals [5, 7, 43–45], particularly if they were ‘only’ coughing [44, 46].

In our sample of networked providers, pharmacies and medicine vendors had higher rates of providing or referring patients for TB testing and treatment, while the private hospitals and clinics provided appropriate care for every patient who visited them. This was due to the support this network of providers received, and the fact that most of the drug stores in our sample were located close to the private clinics and hospitals in the same network. Few studies have looked at the differences in referrals between different types of private providers; however, studies in Nigeria and in similar settings show that case notifications and referrals for TB care from the private sector has been historically very low and in need of intervention [42-44, 47-49]. The SHOPS Plus project, in collaboration with other stakeholders in Nigeria, have implemented several strategies to strengthen linkages between the private sector and the public TB delivery system [50].

Several studies found that individuals with chest symptoms are very likely to minimise symptoms like cough or fever [38, 51–53]. Similar results have been reported in other high-burden TB settings where symptoms like weight loss were associated with faster care-seeking, in comparison to cough and fever which were perceived to be ‘normal’ [7, 51, 52, 54]. In some settings, fever or headaches have been found to shorten delays [55–57].

In charting patient pathways, our results show that all the patients who chose a private hospital (including recruitment facility) as their first or second provider did not use an additional provider. This was likely influenced by the fact that our recruitment facilities, which were supported sites, represented the first facility for 71 (40%) of all participants. We also found a similar pattern among patients who used a private hospital as second provider - of not going to another provider. These findings agree with studies from several high burden TB countries, where hospitals, in public and private sector, missed diagnosing TB patients [49, 58–60]. A pre-covid study in Nigeria found patients having up to 5 pre-NTP provider visits [61]

Qualitative themes related to misdiagnosis, lack of TB testing or referrals to other facilities were also common. Congestion and waiting times in health facilities also discouraged participants from seeking care, but that finding might have been a result of our purposive sampling in high-volume facilities.

While Nigeria has shown remarkable increases in number of annual case notifications [1], there are still approximately 208,000 missing cases estimated annually. Our findings are in concordance with several studies on TB care cascade showing that access to diagnostic services, or Gap 1, is the largest gap in the care cascade [32, 33, 42, 62]. Participants faced several barriers in accessing diagnostic services, especially during the start of the COVID-19 pandemic, and yet the responsibility for being diagnosed rests mostly on the patient, just like before the pandemic [38]. Our findings agree with other studies showing TB positive participants face longer diagnostic delays [32, 33, 38, 63, 64], often due to lack of access to a diagnostic test [32].

### Determinants of private sector use and numbers of providers

Our logistic regressions identified factors influencing private sector use and total number of providers encountered within the context of our study. Our participants were all recruited within a network of dedicated private providers in Kano and Lagos, making them distinct from the general population in terms of their inclination and capacity to utilize private healthcare. Participants in Lagos State were more likely to use the private sector first (93%) before going to the public sector, compared to 67% in Kano. This might be because self-employed individuals might be more flexible with the time required to seek, and the lower cost of public healthcare. Two studies in India found young age, females, higher level of education and income group associated with private sector use [65, 66]. Several studies in Nigeria and other countries have suggested that patients are reluctant to use the public sector because of longer waiting times, lower quality of care, or poor provider attitudes, while on the contrary, private facilities have a reputation of better quality at a higher cost [38, 67, 68].

Patients who had difficulty in breathing as a symptom were more likely to use more than one provider. Several studies have shown that patients with chest symptoms delay care-seeking for a variety of reasons and also contact several private providers before diagnosis [69–72]. This is likely because of the symptom minimization observed in several countries, where patients do not immediately seek hospital care for cough until symptoms deteriorate, preferring to self-medicate, or visit informal providers [38, 46, 72–74]. The use of more than one provider has also been shown to be partly responsible for prolonged delays in TB diagnosis and treatment [5, 75–77].

### Impact of COVID-19

Participants in our study reported that the impact of COVID-19 on care-seeking was mostly felt during the lockdown periods, with a subsequent recovery to pre-pandemic levels. This finding aligns with the quick recovery of TB case notification in Nigeria compared to other countries [1]. Disruptions to TB services have been widely documented in many high-burden settings [28, 78, 79]. Lockdown and the movement restrictions posed barriers to accessing care, leading to delayed care seeking due to difficulties in finding affordable transportation. Other concerns included the affordability of hospital care during the lockdown, fear of being diagnosed with COVID-19 or overburdened public hospitals. Some participants perceived their symptoms to be minor, further contributing to delayed care-seeking. Some participants narrated fearful healthcare workers’ attitudes towards them, particularly during the early days of the pandemic.

From the survey responses, most participants indicated that COVID did not affect their care-seeking beyond the lockdown period. However, the qualitative data showed fear of COVID was a predominant theme resulting in delayed care-seeking, albeit short-lived.

### Limitations

Our study has a few limitations. First, we were unable to calculate total provider delays across all provider encounters as our time measure were categorical and not exact dates. We also did not capture data on the total visits for each provider, as this would have provided a greater understanding of the total number of times participants sought care for their symptoms. Our analysis is limited by non-random purposive sampling for selecting facilities and patients and selection bias. We cannot say whether a study including patients diagnosed in the public sector or outside of the network of providers supported by the SHOPS Plus program will show similar results. There was also no way to determine exactly if our sampled facilities were representative of the distribution of all facilities in Kano and Lagos. Finally, as patients were asked to recall events that had happened earlier, there might have been some recall bias in the responses. We attempted to mitigate this effect by only selecting recently tested patients, with encounters with the facilities dating back no further than January of 2021, or 5 months before our data collection.

### Conclusion

Our patient pathway study in two major cities in Nigeria showed relative shorter individual pathways to care, given the unique context. It emphasizes the importance of involving lower cadres in case identification and referral efforts, irrespective of the sector. Despite well-funded initiatives, delays persist, albeit potentially more manageable. Our study suggests that COVID-19 does not pose a lasting hurdle for the National TB control efforts among patients who are willing and able to utilise private healthcare. More patient education on the importance of getting tested with prolonged coughing, as well as improving referral systems and provider training within the private and public healthcare sectors are needed in our study settings. The WHO calls for multisectoral and intersectoral engagement in the fight against TB, and our findings support evidence from other high-burden settings that private sector provider engagement is critical.

## Data Availability

All data produced in the present study are available upon reasonable request to the authors

## Acknowledgments

We would like to thank the Abts Associates team, particularly Elaine Baruwa and all the field staff and providers that participated in this survey. We thank the COVET study team and the McGill International TB centre, particularly Caroline Vadnais, for sustained support throughout the process of this study. The authors also thank Guy Stallworthy, Sameer Kupta and Kayla Laserson of the Bill and Melinda Gates Foundation for their support.

## Appendix

**Appendix 1:** Concurrent mixed methods and participant recruitment flow chart

|  |  |  |
| --- | --- | --- |
| Facility selection | **Sampling of recruitment facilities (n=18)**<br>1. Divided Lagos and Kano into 3 each by senatorial zones (n=6)<br>2. Selected 3 highest volume facilities based on notifications in the last 3 months from each senatorial zone, ensuring different local government locations (n =18) |  |
| Participant recruitment | **Participant recruitment (337)**<br>1. TB client registers were reviewed starting from the most recent listings (within 6 months)<br>2. Clients over 18 years old and HIV negative with confirmed TB negative (presumptive) and TB positive (confirmed) sputum test results were included<br>3. Out of 337 patients contacted, 212 (63%) consented for the quantitative surveys, and 46 (14%) consented for qualitative interviews, and 77 (23%) were unreachable. Response rate = 77%.<br>4. Surveys were scheduled and conducted until quota (180 surveys) was reached. |  |
| Data collection<br>10<sup>th</sup>- 28<sup>th</sup><br>May 2021 | **Quantitative survey (n=180)**<br>1. In each state - 90 surveys (45 presumptive and 45 confirmed TB)<br>2. Questions on delays and pathways to TB testing and treatment and impact of COVID | **Qualitative interviews (n=20)**<br>1. In each state - 10 in-depth interviews<br>2. Questions explored facilitators and barriers to TB care initiation and impact of COVID |
| Data analysis | Descriptive statistics and logistic regression | Thematic coding |
| Results | **Integration of mixed methods results**<br>1. Statistically significant results were compared with qualitative themes<br>2. Mixed methods results were compared for complementarity and discordance |  |

**Appendix 2:** Quota sample distribution by site

| State | Senatorial Zone | Facility Name | LGA | # of network facilities | Facility Type | Facility registers<br>Jan – May 2021 |  | Surveyed<br>Jun 2021 |  |
| --- | --- | --- | --- | --- | --- | --- | --- | --- | --- |
|  |  |  |  |  |  | TB negative | TB positive | TB negative | TB positive |
| Kano | Kano Central | Zakirai Nursing Home & Maternity | Dala | 61 | Hospital | 187 | 56 | 5 | 5 |
|  | Kano Central | Kura Surgery and Maternity Clinic | Kura | 14 | Hospital | 168 | 52 | 5 | 5 |
|  | Kano Central | International Hospital | Fagge | 122 | Hospital | 447 | 50 | 5 | 5 |
|  | Kano Central | Al-Nouri Hospital | Tarauni | 115 | Hospital | 515 | 31 | 5 | 5 |
|  | Kano North | Yakubu Dannono Memorial Hospital | Tofa | 11 | Hospital | 318 | 39 | 5 | 5 |
|  | Kano North | Dawanau Clinic & Maternity | Dawakin Tofa | 7 | Hospital | 152 | 3 | 5 | 5* |
|  | Kano South | Al-Hilal Nursing & Maternity Home | Wudil | 48 | Nursing Home | 244 | 69 | 5 | 5 |
|  | Kano South | AHIP FHC Clinic | Garko | 29 | Hospital | 566 | 81 | 5 | 5 |
|  | Kano South | H. Nasiha Clinic and Maternity Centre | Gaya | 45 | Hospital | 449 | 103 | 5 | 5 |
| Lagos | Lagos Central | St. Lukes Hospital | Lagos Mainland | 37 | Hospital | 75 | 14 | 5 | 5 |
|  | Lagos Central | Fenhitola Royal Consult | Lagos Island | 63 | Hospital | 156 | 14 | 5 | 5 |
|  | Lagos Central | St. Kizito Clinic Lekki | Eti-Osa | 30 | Hospital | 591 | 84 | 5 | 5 |
|  | Lagos East | Eloka Nursing Home | Ikorodu | 72 | Nursing Home | 100 | 24 | 5 | 5 |
|  | Lagos East | Al Sadiq Hospital | Kosofe | 46 | Hospital | 71 | 18 | 5 | 5 |
|  | Lagos East | R-Jolad Hospital | Shomolu | 27 | Hospital | 179 | 12 | 5 | 5 |
|  | Lagos West | St. Theresa Hospital | Ajeromi-Ifelodun | 113 | Hospital | 517 | 89 | 5 | 5 |
|  | Lagos West | Medservice Hospital | Alimosho | 335 | Hospital | 58 | 23 | 5 | 5 |
|  | Lagos West | St Catherine Med. | Oshodi-Isolo | 70 | Hospital | 115 | 19 | 5 | 5 |
| * Sampled patients included 2 patients who were diagnosed within recruitment period earlier to complete quota |  |  |  |  |  |  |  |  |  |

**Appendix 3:** Characteristics of respondents and non-respondents

|  |  | Patient that consented<br>N=180 | Patient that declined<br>N=80 | Overall<br>N=260 |
| --- | --- | --- | --- | --- |
| **State** | Kano | 90 (50.0%) | 50 (62.5%) | 140 (53.8%) |
|  | Lagos | 90 (50.0%) | 30 (37.5%) | 120 (46.2%) |
| **Gender** | Female | 70 (38.9%) | 30 (37.5%) | 100 (38.5%) |
|  | Male | 110 (61.1%) | 50 (62.5%) | 160 (61.5%) |
| **Age Group** | 18-24 | 24 (13.3%) | 12 (15.0%) | 36 (13.8%) |
|  | 25-34 | 63 (35.0%) | 24 (30.0%) | 87 (33.5%) |
|  | 35-44 | 50 (27.8%) | 22 (27.5%) | 72 (27.7%) |
|  | 45-54 | 21 (11.7%) | 7 (8.8%) | 28 (10.8%) |
|  | 55+ | 22 (12.2%) | 15 (18.8%) | 37 (14.2%) |
| **TB Status** | TB negative | 90 (50.0%) | 43 (53.8%) | 133 (51.2%) |
|  | TB positive | 90 (50.0%) | 33 (41.2%) | 123 (47.3%) |
|  | Missing | 0 (0%) | 4 (5.0%) | 4 (1.5%) |

**Appendix 4:** Average number of encounters by type of provider seen first

| Initial Provider contact | Total number of individuals | Mean Number of Encounters (SD) | Median (Min, Max) |
| --- | --- | --- | --- |
| Private hospital or nursing home | 79 | 1.10 (0.304) | 1.00 [1.00, 2.00] |
| Community Pharmacy or Medicine Vendor | 57 | 2.35 (0.612) | 2.00 [2.00, 5.00] |
| Public Hospital | 26 | 2.31 (0.618) | 2.00 [2.00, 4.00] |
| Public outpatient clinic | 6 | 3.00 (1.265) | 2.50 [2.00, 5.00] |
| Private laboratory | 4 | 2.00 (0.00) | 2.00 [2.00, 2.00] |
| Community Health Worker/CHEW | 3 | 2.67 (0.577) | 3.00 [2.00, 3.00] |
| Private outpatient clinic, health centre, or dispensary | 3 | 3.33 (0.577) | 3.00 [3.00, 4.00] |
| Traditional healers | 1 | 3.00 (NA) | 3.00 [3.00, 3.00] |
| Other, specify | 1 | 4.00 (NA) | 4.00 [4.00, 4.00] |
| TOTAL | 180 | 1.84 (0.864) | 2.00 [1.00, 5.00] |

**Appendix 5:** Selected qualitative themes and example quotes

| Theme # | Theme Description | Codes | Sample Quote |
| --- | --- | --- | --- |
| 1 | Delays | Self-medication | *"Yes, it was up to a year, it was a gradual thing. I treated typhoid and malaria because that has always been the major sickness I usually have and when I treat, it subsides, and I'll think I'm okay until it crawls back again,"* (#6, Female, TB +, Lagos). |
|  |  | Self-medication/<br>Symptom minimization | *"I just thought it's the normal cough caused by [cold] and which makes me to be taking different cough syrups to be better, but it didn't work"* (#10, Female, TB -, Kano). |
|  |  | Symptom minimization | *"...for instance, you can't just get up and visit the doctor in a haste because of headache. It is just like one going to the farm and feeling the hot sun but knowing that in a matter of time you will feel relief from the effect of the sun. And that was why I spent some few days before visiting the chemist."* (#5, Male, TB +, Kano). |
|  |  | Symptom minimization<br>/Misdiagnosis | *"I was coughing blood. I didn't take it seriously ... when I came home, my dad asked me to take herbs ... it persisted, so my mum took me to the chemist who gave me drugs to take for 10 days. I felt if I completed the 10 days it would stop. After completing it, it didn't stop,"* (Male, TB Positive, Lagos).<br>(#9, Male, TB +, Lagos). |
|  |  | Misdiagnosis | *"...Then we didn't know what the sickness was, we didn't know what type of cough it was. I went to chemist, I was tested and given some drugs for catarrh and I felt relieve after taking it but whenever the medicine is finished the cough will be back"* (Female, TB Positive, Kano).<br>(#7, Female, TB +, Kano). |
|  |  | Same day treatment | *"I was diagnosed at that time ... in the hospital when we came back ...it was all done here, after the test and checking, drugs was given to us on the same day. ..."* (Female, TB Positive, Kano).<br>(#4, Female, TB +, Kano). |
|  |  | Same day treatment | *"I was told to come back the same day for the test result ... When I came back, I was told about the TB and that I needed to commence treatment. I was asked some questions... how many people I stayed with, if where I stayed had ventilation .... I was given the drugs to take ... When I started treatment, my chest pain started reducing"* (#9, Male, TB +, Lagos).<br>(#9, Male, TB +, Lagos). |
| 2 | Pathways | Symptoms progressing/<br>misdiagnosis | *"when it started, I thought, it was just a mild cough, that was why I used cough syrup. It was later, when I started coughing out blood, that I now thought that this has passed normal cough because it was so serious, that was why I went to the hospital, they now referred me [from one public hospital to another] and from there, we know came here [private hospital]"* (Male, TB positive, Lagos).<br>(#5, Male, TB +, Lagos). |
|  |  | Symptoms progressing | *"...it was excessive coughing that had been disturbing me; and I use to go and collect drugs ... but that didn't change anything, so I took drugs for a long time"* (Male, TB positive, Kano).<br>(#2, Male, TB +, Kano). |
| 3 | Choice of initial provider | Proximity and convenience | *"The chemist is closest to us, without spending on transportation or ... other services. Chemist services are daily and constant compared to the hospital where you will be given time to see the doctors and they rarely operate at night."* (#10, Female, TB -, Kano).<br>(#10, Female, TB -, Kano). |
|  |  | Recommendations of family/friend | *"The whole process took long because there was no sign of cough just that it was eating me ... So, ... when I travelled home, a matron from my family ... asked me if I'd gone for test. I told her I had gone for series of tests and she told me to go for TB test which made me laugh and I told her ...I clearly wasn't coughing. ... she told me it doesn't matter because TB has a way of manifesting by eating one up internally .... When I got back to Lagos, someone else told me the same thing which made me go for test at the Lab... Not until when I came here and the doctor prescribed 3 tests which I did and that was when I started coughing which was increasing gradually ... up to a week and at a point I started coughing out blood".*<br>(#6, Female, TB +, Lagos). |
| 4 | Use of multiple providers | Misdiagnosis, recommendation of friend | *"I went to the chemist [after my symptoms started] ... to a private doctor for cough syrup and tablets. Then ...to a public hospital during pandemic, there was long queue...I took herbs... went to another hospital. This boy that works in a hospital asked if I had conducted a test ... he then said I should go to the lab with a note he has written. " (#5, Male, TB +, Kano).*<br>*(#5, Male, TB +, Kano).* |
|  |  | Misdiagnosis, recommendation of friend | *"It has been the long I had the symptoms. My mum, we were thinking that it was malaria or fever, so I've been using malaria drugs since February, because our family nurse told us that it was malaria, so she was just giving me injections, and drugs and I did not like it. We went to the hospital by March and they told me that it was fever, so I was also given injections, and drugs ... I told them that I was coughing and having chest pain, but the drugs they gave me... were just for malaria. I was complaining to the doctor ...that I was coughing. I did blood tests too but the drugs they were giving me, it was actually not relieving, so I told my doctor in our church... It was him that referred me to ... to go for a x-ray...because my mum once had TB, maybe it could be TB, he guessed... I went for x-ray...and it was TB and the sputum test came out too, it was TB".*<br>*(#1, Female, TB +, Lagos).* |
| 5 | Impact of COVID | Fear of diagnosis | *I will be thinking that if I have this malaria, it's better I go to the chemist to buy the anti-malaria drug than going to the hospital where I will be told that I have COVID-19 just because I have malaria symptoms.*<br>*(Female TB positive, Lagos)*<br>*(#1, Female, TB +, Lagos).* |
|  |  | Provider attitude, overburdened facilities, and infection control mandates | *"Actually, there are a lot of differences you will notice. During the COVID-19 period, the doctors around will be harassing people without considering their condition. ... And those COVID-19 protocols are some of the serious challenges we faced. At that time, they will tell you to use face mask, use hand sanitizer and social distancing... And there was congestion, too much death rate. Actually COVID-19 has affected my access to health care services a lot".*<br>*(#10, Female, TB -, Kano).* |
|  |  | Movement restrictions, infection control mandates, provider attitude | *"As villagers, we don't know much about COVID-19. It ... affected me during lockdown that I couldn't get access to the hospital. My major challenge was during the lockdown. I don't like that covid-19 protocols, wearing of face masks does not allow me to breath well, that social distancing. And above all even the healthcare providers are afraid to attend to us because COVID-19 and TB are communicable diseases and have similar symptoms".* |
|  |  |  | (#9, Male, TB +, Kano). |

